# Menopausal symptom burden as a predictor of mid- to late-life cognitive function and mild behavioral impairment symptoms: A CAN-PROTECT study

**DOI:** 10.1101/2024.03.13.24304247

**Authors:** Jasper F.E. Crockford, Dylan X Guan, Gillian Einstein, Clive Ballard, Bryon Creese, Anne Corbett, Ellie Pickering, Adam Bloomfield, Pamela Roach, Eric E Smith, Zahinoor Ismail

## Abstract

**Background:** Recent evidence suggests the experience of menopausal symptoms (i.e., perimenopausal symptoms) may be associated with cognitive and behavioural changes. We investigated these two relationships in a sample of post-menopausal females.

**Design:** Cross-sectional observational study.

**Setting:** Participant data was collected from the Canadian Platform for Research Online to Investigate Health, Quality of Life, Cognition, Behaviour, Function, and Caregiving in Aging (CAN-PROTECT) study.

**Participants:** 896 post-menopausal female participants.

**Methods:** Menopausal symptom burden was operationalized by summing the total number of recalled perimenopausal symptoms experienced. Cognitive function was measured using the Everyday Cognition (ECog-II) Scale, with higher scores reflecting greater severity. Mild Behavioral Impairment (MBI) was measured using the Mild Behavioral Impairment Checklist (MBI-C), with higher scores reflecting greater severity. A negative-binomial regression model examined the relationship between menopausal symptom burden and cognitive function, while a zero-inflated negative binomial regression model examined the relationship between menopausal symptom burden and MBI symptoms. Models adjusted for age, years of education, age of menopausal onset, type of menopause, and hormone therapy (HT). Age of menopausal onset and use of HT in the two associations were investigated with moderation analyses.

**Results:** Greater menopausal symptom burden was associated with higher ECog-II total scores (b [95% confidence interval (CI)] = 5.37 [2.85, 7.97]) and higher MBI-C total scores (b [95% CI] = 6.09 [2.50, 9.80]). Use of HT did not significantly associate with ECog-II total scores (b [95% CI] = -10.98 [-25.33, 6.35]), however, HT was significantly associated with lower MBI-C total scores (b [95% CI] = -26.90 [-43.35, -5.67]).

**Conclusions:** Menopausal symptom burden is associated with poorer cognitive function and more MBI symptoms in mid- to late life. HT may help mitigate symptoms of MBI. These findings suggest the experience of menopause may indicate susceptibility to cognitive and behavioural changes, both markers of dementia.

## Introduction

As the number of older adults continues to rise, those living with Alzheimer’s disease (AD) and related dementias globally are estimated to increase three-fold from 50 million in 2015 to 152 million by 2050^1^. Females (i.e., assigned female sex at birth) have a three-fold greater risk of developing AD^2^, and will be disproportionately affected by this increasing global dementia burden^3^. As prevalence of dementia increases with age^4^, the primary explanation offered for the sex discrepancy is greater longevity in females^5^. However, whether longevity alone explains this sex difference remains uncertain; extant literature has proposed there are unique factors that may increase female risk^4^. One proposed factor that may confer special risk to females is the loss of estradiol at menopause.

Menopause describes the permanent cessation of menses^6^. There are “many menopauses”, each with their own cognitive sequelae^7^, which can occur either spontaneously (i.e., naturally) or due to medical or surgical reasons (e.g., cancer treatments, oophorectomy, hysterectomy) in mid-adulthood^8^. Menopause comprises physiological, cognitive, and behavioural symptoms, likely related to declining 17-b-estradiol (estradiol) levels – the most common type of estrogen^9^. Menopausal symptoms (e.g., poor sleep, depression, etc.) may themselves create risk of later brain pathology, such as the development of AD-related neuropathologies^10–13^.

Later-life decline in cognitive function can signal risk of incident dementia^14,15^ and menopause is associated with cognitive changes ^7,8,16,17^. Recent cross-sectional studies have found spontaneous menopause to be associated with poorer cognitive performance in the domains of language^18,19^, memory^8,20–23^, visuo-spatial^24^, and executive function^20,24–28^. However, studies have not determined the nature of this association — whether cognition is related to ovarian cessation, itself, or to an additive effect of multiple menopausal symptoms. Longitudinally, earlier entry into menopause, whether spontaneously or surgically, has been linked to greater risk of cognitive impairment, AD, and other dementias^16,17,29–31^. Hormone therapies (HT) composed of estrogen variations and hormonal contraceptives, including oral contraceptive, have long been used to ameliorate menopausal symptoms. However, whether HT use is associated with improvement in cognition or reduced incidence of dementia remains unclear^8,32–35^.

Analogous to cognitive changes, later-life behavioural changes can also signal risk for incident dementia. The International Society to Advance Alzheimer’s Research and Treatment’s criteria for the syndrome Mild Behavioral Impairment (MBI)^36^ operationalizes risk by identifying behavioural and neuropsychiatric symptoms (NPS) that are later-life emergent and persistent. NPS meeting these criteria confer significantly higher risk of incident cognitive decline and dementia compared to conventionally measured behavioural and psychiatric symptomatology^37–47^ However, unlike the relationship between ovarian cessation (i.e., any type of menopause) and cognition, evidence is sparse on the association between ovarian cessation and MBI. This evidence gap warrants further investigation.

We investigated the relationship between menopausal symptoms and later-life changes in cognitive function and behaviour; both risk factors for dementia, which may also represent preclinical/prodromal AD^14,15,42,43,48–55^. We hypothesized that individuals who experienced a greater number of menopausal symptoms (i.e., greater menopausal symptom burden) would subsequently experience poorer cognitive function and greater MBI symptom burden compared to individuals who experienced fewer menopausal symptoms.

## Methods

### CAN-PROTECT study

Data were collected from the ongoing Canadian Platform for Research Online to Investigate Health, Quality of Life, Cognition, Behaviour, Function, and Caregiving in Aging (CAN-PROTECT) study^56^. CAN-PROTECT is a Canada-wide online observational cohort study of brain aging that seeks to assess the roles of demographic, medical, environmental, and lifestyle factors on risk and resilience. To participate in CAN-PROTECT, individuals must be dementia-free residents of Canada, aged ≥18 years, with access to an internet-connected computer or tablet. In CAN-PROTECT, participants complete a detailed demographics questionnaire, and an annual series of mandatory and optional online assessments of cognition, behaviour, function, health, wellness, quality of life, medical/psychiatric history, and lifestyle. CAN-PROTECT was approved by the Conjoint Health Research Ethics Board at the University of Calgary and began participant recruitment on March 8, 2024. Informed consent was obtained from all participants at registration.

### Participants

Baseline participant responses pertaining to demographics, cognition, and emotional/psychiatric symptoms were analyzed, in addition to the optional fertility and menopause questionnaire. From the initial sample of 1984 participants (77.7% female), 896 (100% female) who reported being post-menopausal were included here. Of these, 667 had spontaneous menopause, 162 menopause due to hysterectomy/oophorectomy, and 67 menopause due to other medical reasons (Figure 1).

**Figure 1.**
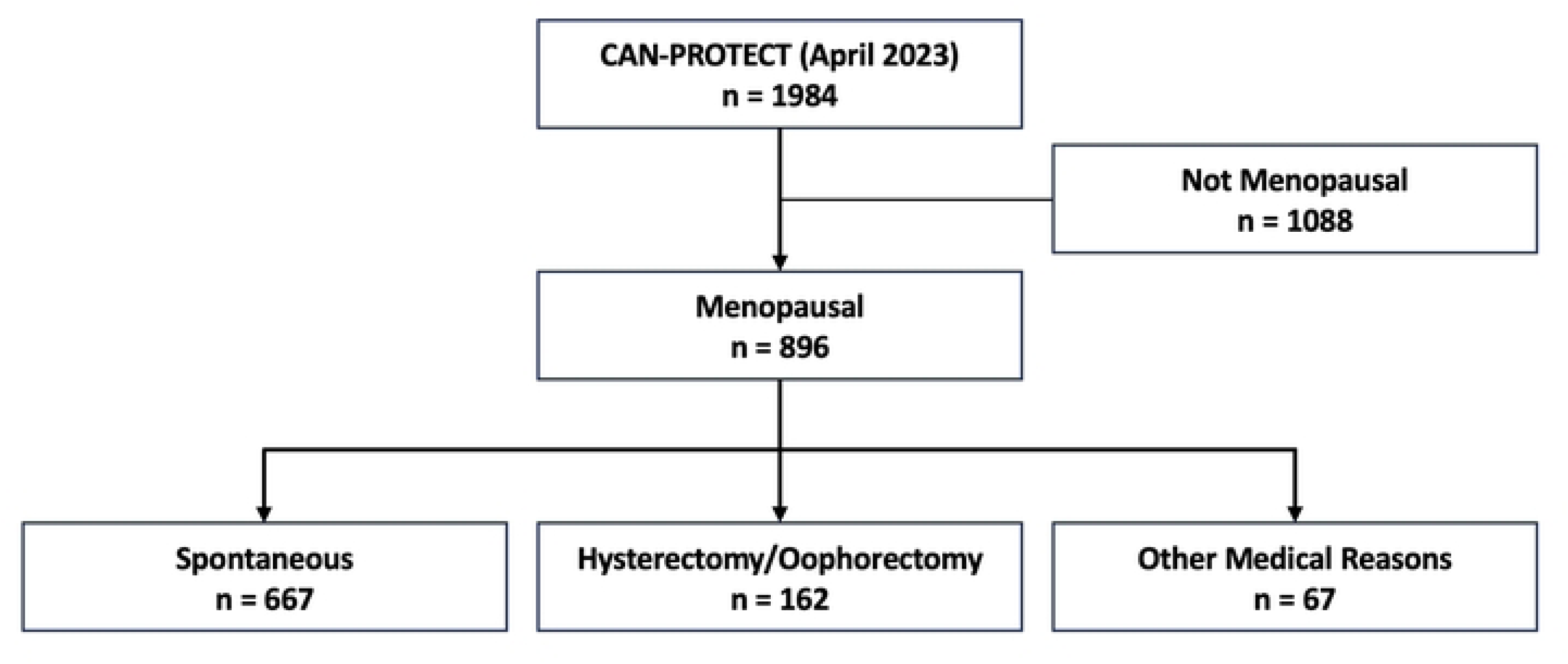
Flowchart of participants from CAN-PROTECT included for analysis. Abbreviations: CAN-PROTECT, Canadian Platform for Research Online to Investigate Health, Quality of Life, Cognition, Behaviour, Function, and Caregiving in Aging.

### Measurements

#### Menopausal symptom burden scale

The recalled presence or absence of 11 perimenopausal symptoms reflecting vasomotor, cognitive, and behavioural/neuropsychiatric symptoms was assessed using the fertility and menopause questionnaire. The scale comprised 11 symptoms including irregular periods, hot flashes, chills, vaginal dryness, weight gain, slowed metabolism, night sweats, sleep problems, mood symptoms, inattention or forgetfulness, and other unnamed symptoms. Total menopausal symptom burden was calculated by summing the number of self-reported menopausal symptoms experienced by participants. Menopausal symptom burden scores ranged from 0 to 11, with 0 representing the least burden (i.e., no symptoms present) and 11 representing the greatest burden (i.e., presence of all 11 symptoms).

#### Everyday cognition scale

Cognitive function was measured with the revised Everyday Cognition (ECog-II) Scale^57^, developed to identify new onset cognitive symptoms in dementia at-risk populations. The scale consists of 41 questions that assess changes in memory (9 items), language (8 items), visual-spatial and perceptual abilities (8 items), planning (5 items), organization (4 items), and executive function (4 items). Questions are rated from 0-3 (0 = no change, 1 = occasionally worse, 2 = consistently a little worse, 3 = much worse) relative to a participant’s own baseline, with higher scores reflecting more consistent impairment. Total ECog-II scores were calculated by summing all item scores. Thus, a higher ECog-II total score indicate worse cognitive function.

#### Mild Behavioral Impairment checklist

MBI symptoms were assessed using the Mild Behavioral Impairment Checklist (MBI-C)^58,59^, developed to identify new onset behavioural symptoms in dementia at-risk populations. The MBI-C consists of 34 items that evaluate later-life emergent and persistent neuropsychiatric symptoms in five domains: apathy (6 items), affect (6 items), impulse dyscontrol (12 items), social cognition (5 items), and psychosis (5 items). Like the ECog-II, MBI-C items are scored from 0-3, with higher scores reflecting greater symptom change from baseline. A total MBI symptom burden score was calculated by summing all item responses to create a total out of 102. Higher MBI-C total scores reflected greater MBI symptom burden.

### Statistical analysis

Participant characteristics were summarized using descriptive statistics (count, percentages, mean, and standard deviation). The data were overdispersed counts (i.e., variance>mean). Thus, to assess the relationship between menopausal symptom burden and cognitive function, a negative binomial regression model was used. Menopausal symptom burden was treated as the exposure variable and ECog-II total score as the outcome variable.

Similarly, to assess the relationship between menopausal symptom burden and MBI symptoms, a zero-inflated negative binomial regression model was used. Menopausal symptom burden was treated as the exposure variable and MBI-C total score as the outcome variable. Both models were adjusted for age, total years of education, type of menopause, age of menopausal onset, and use of HT. Type of menopause consisted of two conditions: 1) spontaneous; and 2) due to surgical or medical reasons. Additionally, the use of HT consisted of three conditions: 1) no HT use; 2) use of progestin or unnamed HT forms; and 3) and use of HT. In both models, two interaction terms were included: 1) age of menopausal onset (participant age of menopausal onset was compared to the mean age of menopausal onset); and 2) use of HT.

## Results

Of the 896 included participants assigned female sex at birth, 895 identified as women, while one participant identified as two-spirit. The mean (±standard deviation; SD) age of participants was 64.2±7.3 years with 15.5±5.0 years of education. Participants reported the onset of menopause at a mean age of 49.4±6.5 years; spontaneous menopause, menopause due to hysterectomy/oophorectomy, and menopause due to other medical reasons occurred at the mean ages of 51.3±4.2, 42.0±8.7, and 47.3±5.2 years, respectively. A total of 666 (74.3%) participants experienced menopausal symptoms. To manage perimenopausal symptoms 166 (24.9%) participants used HT, including estradiol-based therapy (n = 41, 6.2%), conjugated estrogens (n = 34, 5.1%), estrogens-progesterone combinations (n = 73, 11.0%), or estrogens-progestin combinations (n = 18, 2.7%). Twelve participants reported the use of progestin (1.8%) and 47 reported the use of unnamed HT types (7.1%) (see Table 1 for details of participant demographics).

**Table 1.** Demographic characteristics of participants.

| Demographic Characteristic | n(%) | M(SD) | Range(years - years) |
| --- | --- | --- | --- |
| Sex at Birth |  | --- | --- |
| *Female* | 896(100%) | --- | --- |
| *Male* | 0(0%) | --- | --- |
| Identifies As |  |  |  |
| *Female* | 895(99.9%) | --- | --- |
| *Male* | 0(0%) | --- | --- |
| *Two-Spirit* | 1(0.1%) | --- | --- |
| *Non-Binary* | 0(0.0%) | --- | --- |
| *Other* | 0(0.0%) | --- | --- |
| Age |  | 64.2(7.3) | 41-88 |
| Total Years of Education | --- | 15.5(5.0) | --- |
| Age of Menopausal Onset | --- | 49.4(6.5) | 12-62 |
| Type of Menopause |  |  |  |
| *Spontaneous* | 667(74.4%) | 51.3(4.2) | 12-62 |
| *Hysterectomy and/or* | 162(18.1%) | 42.0(8.7) | 12-62 |
| *Oophorectomy* |  |  |  |
| *Other Medical Reasons* | 67(7.5%) | 47.5(5.2) | 30-58 |
| Experience of Perimenopausal | 666(74.3%) | --- | --- |
| Symptoms |  |  |  |
| HT Use | 166(24.9%) | --- | --- |
| *Estradiol* | 41(6.2%) | --- | --- |
| *Conjugated Estrogens* | 34(5.1%) | --- | --- |
| *Combination Estrogen-* | 73(11.0%) | --- | --- |
| *Progesterone* |  |  |  |
| *Combination Estrogen-* | 18(2.7%) | --- | --- |
| *Progestin* |  |  |  |
| Progestin or Unnamed HT Use |  |  |  |
| *Progestin* | 12(1.8%) | --- | --- |
| *Unnamed HT* | 47(7.1%) | --- | --- |
| Origin |  |  |  |
| *North American* | 444(49.6%) | --- | --- |
| *First Nation* | 17(3.8%) | --- | --- |
| *Metis* | 12(2.7%) | --- | --- |
| *Inuit* | 0(0.0%) | --- | --- |
| *European* | 764(85.3%) | --- | --- |
| *Caribbean* | 7(0.8%) | --- | --- |
| *South American* | 6(0.7%) | --- | --- |
| *African* | 7(0.8%) | --- | --- |
| *Asian* | 18(2.0%) | --- | --- |
| *Oceania* | 3(0.3%) | --- | --- |

Abbreviations: HT, hormone therapies

From the list of 11 menopausal symptoms, participants endorsed a range of 0 to 10 (see Figure 2) and reported a mean menopausal symptom burden of 3.7±2.8 (spontaneous = 3.8±2.8, due to surgical or medical reasons = 3.5±2.8). The most frequently endorsed perimenopausal symptom was hot flashes (n = 583, 88%) followed by night sweats (n = 496, 70%). The least endorsed perimenopausal symptoms were chills (n = 92, 14%) and other symptoms not listed (n = 31, 5%). Frequency comparison of participant endorsed and not endorsed perimenopausal symptoms, reported from the CAN-PROTECT fertility and menopause questionnaire. Red bars indicate the number of participants who endorsed a perimenopausal symptom, while grey bars indicate the number of participants who did not.

**Figure 2.**
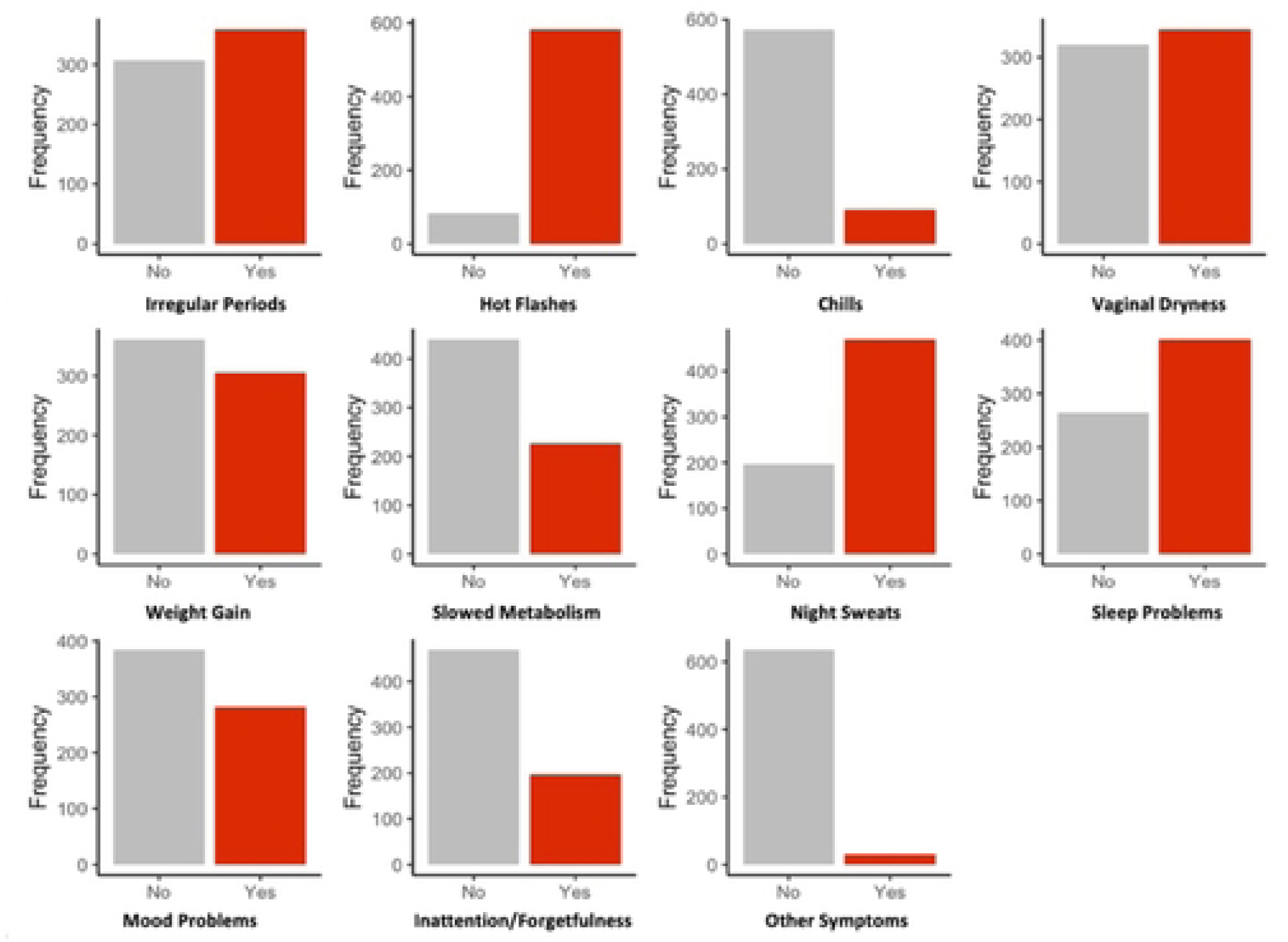
Reported perimenopausal symptoms.

### Everyday cognition and Mild Behavioral Impairment

Collectively, participants reported a mean ECog-II total score of 11.1±10.5, with a range of 0 to 88. Mean MBI-C total symptom burden score was 5.4±7.6, with a range of 0 to 65.

### Menopausal symptom burden and cognitive function

Menopausal symptom burden was associated with poorer cognitive function, according to the results of the negative binomial regression model (see Table 2 and Figure 3). Every additional menopausal symptom was associated with a 5.37% higher ECog-II total score, indicating more consistent symptoms of cognitive dysfunction (95%CI[2.85, 7.97], p<.001). Neither the type of menopause (b=2.78%, 95%CI[-13.10, 21.91], p=.75) nor the age of menopausal onset (b=-1.05%, 95%CI[-2.13, 0.013], p=.068) was significantly associated with ECog-II total score. Similarly, neither HT (b=-10.98%, 95%CI[-25.33, 6.54], p=.20) nor use of progestin or unnamed HT (b=16.93%, 95%CI[-10.93, 56.13], p=.27) were significantly associated with ECog-II total score. Negative-binomial regression coefficient plot of the relationship between menopausal symptom burden (predictor variable) and ECog-II total score (outcome variable). Lines indicate the 95% confidence intervals for the estimate. Predictor variables whose confidence intervals cross the reference line at 1.00 are not significant. Significant predictor variables are indicated with an asterisk.

**Figure 3.**
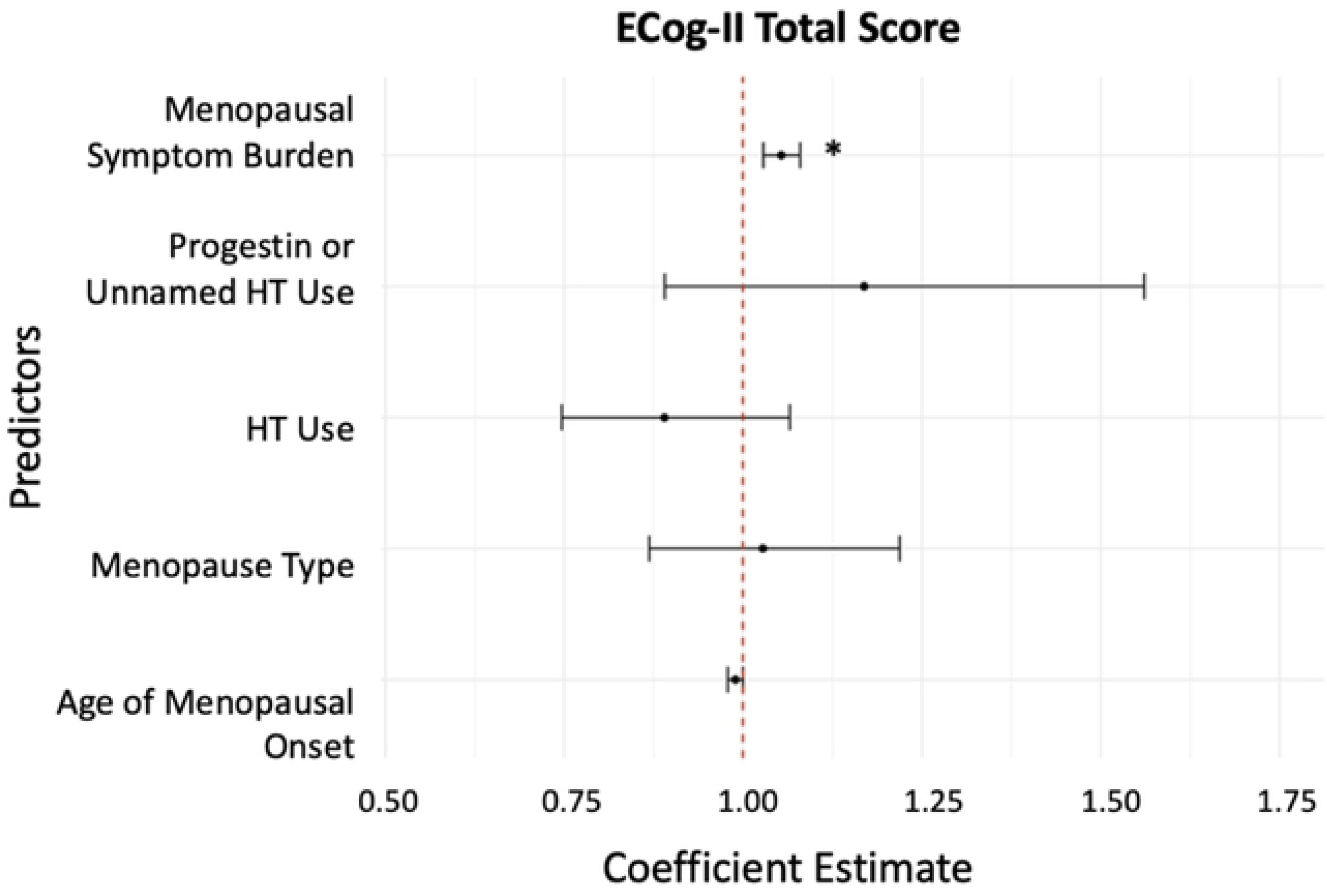
ECog-II total score associations.

**Table 2.**
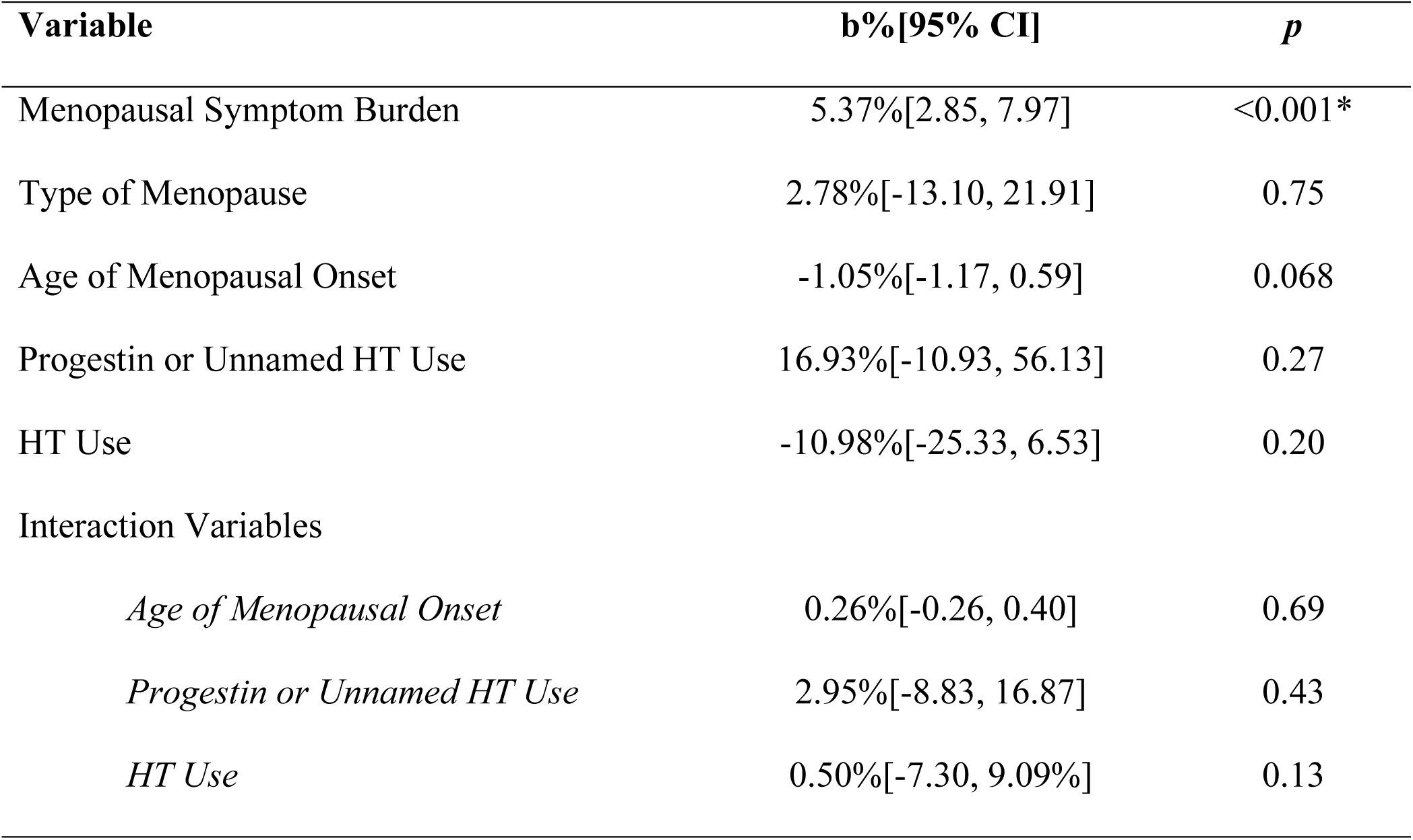
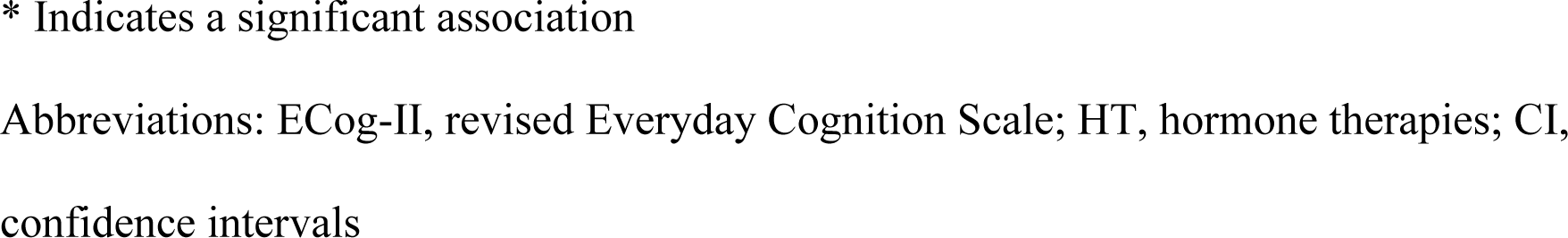
ECog-II total score associations.

| Variable | b%[95% CI] | p |
| --- | --- | --- |
| Menopausal Symptom Burden | 5.37%[2.85, 7.97] | <0.001* |
| Type of Menopause | 2.78%[-13.10, 21.91] | 0.75 |
| Age of Menopausal Onset | -1.05%[-1.17, 0.59] | 0.068 |
| Progestin or Unnamed HT Use | 16.93%[-10.93, 56.13] | 0.27 |
| HT Use | -10.98%[-25.33, 6.53] | 0.20 |
| Interaction Variables |  |  |
| *Age of Menopausal Onset* | 0.26%[-0.26, 0.40] | 0.69 |
| *Progestin or Unnamed HT Use* | 2.95%[-8.83, 16.87] | 0.43 |
| *HT Use* | 0.50%[-7.30, 9.09%] | 0.13 |

\* Indicates a significant association
Abbreviations: ECog-II, revised Everyday Cognition Scale; HT, hormone therapies; CI, confidence intervals

None of age of menopausal onset (b=0.070%, 95%CI[-0.26, 0.40], p=.69), use of HT (b=0.50%, 95%CI[-7.30, 9.09], p=.13), or use of progestin or unnamed HT (b=2.95%,95%CI[- 8.83, 16.87], p=.48) moderated the relationship between menopausal symptom burden and ECog-II total score.

### Menopausal symptom burden and Mild Behavioral Impairment

Menopausal symptom burden was associated with greater MBI symptom burden,f according to the results of the zero-inflated negative binomial regression (see Table 3 and Figure 4). Every additional menopausal symptom was associated with a 6.09% higher MBI-C total score (95%CI[2.50, 9.80], p<.001), indicating more consistent symptoms of MBI. HT use was associated with a 26.90% lower MBI-C total score (95%CI[-43.35, -5.67], p<.016). None of type of menopause (b=4.56%, 95%CI[-18.17, 33.60], p=.72), age of menopausal onset (b=-0.83%, 95%CI[-2.36, 0.74], p=.30), or use of progestin or unnamed HT (b=-19.12%, 95%CI[-44.60, 18.07], p=.27) were significantly associated with MBI-C total score. Zero-inflated negative-binomial regression coefficient plot of the relationship between menopausal symptom burden (predictor variable) and MBI-C total score (outcome variable). The count model coefficient estimates are indicated by the conditional title, while the zero-inflated coefficient estimates are indicated by the zero-inflated title. Lines indicate the 95% confidence intervals for the estimate. Predictor variables whose confidence intervals cross the reference line at 1.00 are not significant and indicated by the colour red. Significant predictor variables are indicated by the colour blue.

**Figure 4.**
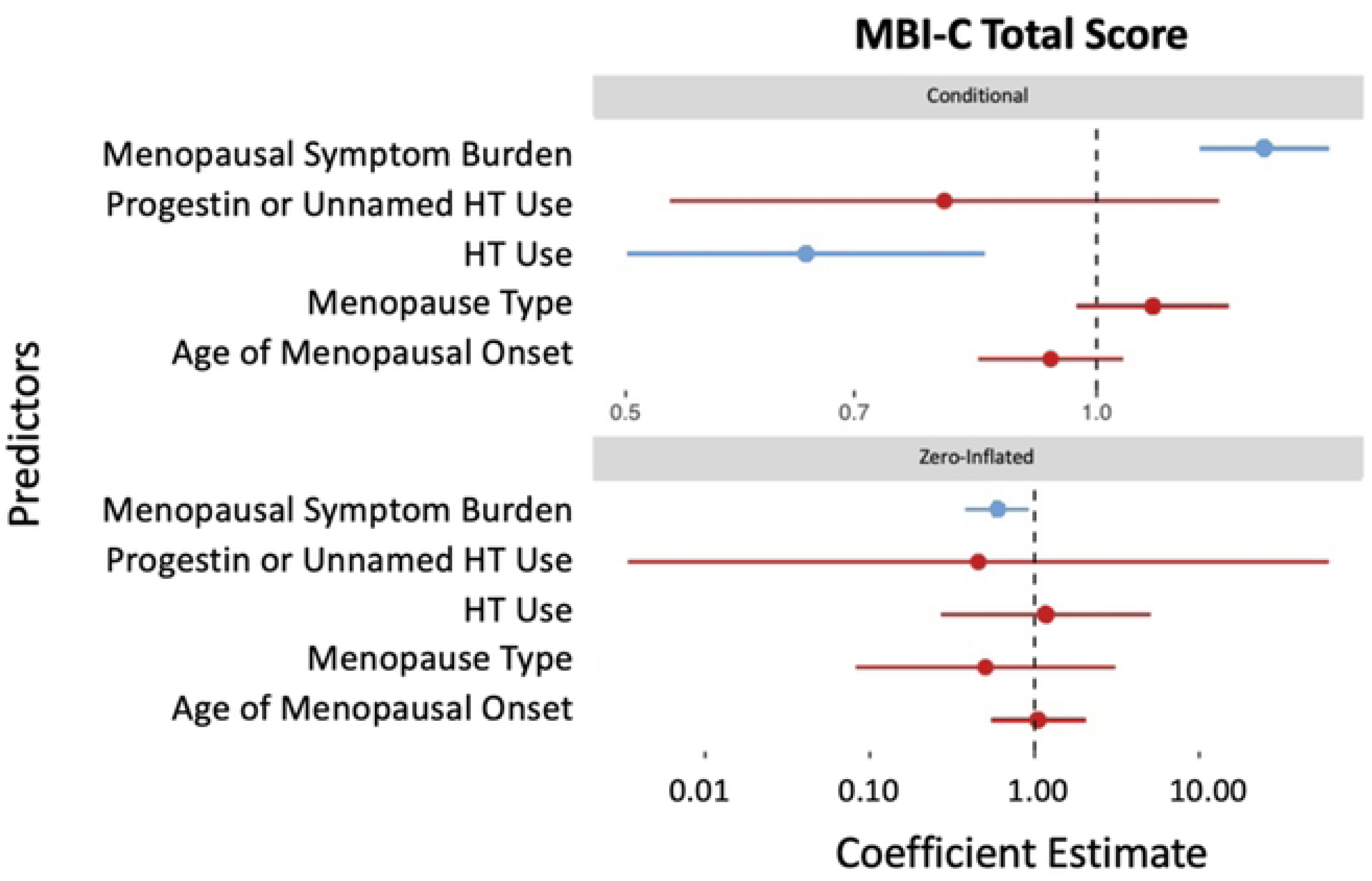
MBI-C total symptom score associations.

**Table 3.** MBI-C total score associations.

| Variable | b%[95%CI] | p |
| --- | --- | --- |
| Menopausal Symptom Burden | 6.09%[2.50, 9.80] | <0.001* |
| Type of Menopause | 4.56%[-18.17, 33.60] | 0.72 |
| Age of Menopausal Onset | -0.83%[-2.36, 0.74] | 0.30 |
| Progestin or Unnamed HT Use | -19.12%[-44.60, 18.07] | 0.27 |
| HT Use | -26.90%[-43.35, -5.67] | 0.016* |
| Interaction Variables |  |  |
| *Age of Menopausal Onset* | -0.066%[-0.50, 0.37] | 0.77 |
| *Progestin or Unnamed HT Use* | 7.22%[-8.01, 25.63] | 0.37 |
| *HT Use* | -0.20%[-11.21, 12.17] | 0.97 |

\* Indicates a significant association
Abbreviations: MBI-C, Mild Behavioral Impairment Checklist; HT, hormone therapies; CI, confidence intervals

Despite the main effect of HT use and lower MBI-C total score, use of HT did not moderate the relationship between menopausal symptom burden and MBI-C total score (b=- 0.20%, 95%CI[-11.21, 12.17], p=.97). Similarly, neither the age of menopausal onset (b=0.066%, 95%CI[-0.50, 0.37], p=.77) nor use of progestin or unnamed HT (b= 7.22%, 95%CI[-8.01, 25.63], p=.37) significantly moderated the relationship between menopausal symptom burden and MBI-C total score.

## Discussion

The experience of menopause can include persistent changes in cognitive function and behaviour, which may extend into later life and identify those at greater risk for incident dementia. In this study, we showed that greater menopausal symptom burden was associated with significantly poorer cognitive function and greater MBI symptom burden, adjusted for age, education, age of menopausal onset, type of menopause, use of HT, and use of progestin or unnamed HT. While use of either HT or progestins or unnamed HT was not significantly associated with changes in cognitive function, use of HT was associated with significantly fewer MBI symptoms. Subsequent analyses revealed that age of menopausal onset and use of HT or progestin or unnamed HT did not moderate the observed associations between menopausal symptom burden and cognition or MBI.

Changes in cognitive function are a defining feature of the normal aging process in females and males^60^. However, Alzheimer disease starts long before diagnosis and understanding the earliest factors influencing its progression is critical. Therefore, identifying later-life emergent and persistent changes in cognitive function may aid in assessing risk for dementia. Other studies have shown that select perimenopausal symptoms may be associated with cognitive changes. For instance, evidence from cross-sectional studies have found perimenopausal females who experienced specific symptoms were more likely to report poorer cognitive function in the domains of attention^24,26^, working memory^8,23^, verbal memory^18,21,24^, visuo-spatial skills^24^, and processing speed^26,27^. However, whether the additive effect of experiencing more than one menopausal symptom is related to later-life changes in cognitive function had yet to be determined. Our first analysis suggests that the degree of menopausal symptom burden (i.e., greater number of menopausal symptoms) is indeed associated with poorer mid-to-later life cognitive function. Thus, our findings are novel in that they show not only are specific menopausal symptoms related to changes in cognitive function, but the overall number of menopausal symptoms experienced are important as well.

Few studies have investigated the association between multiple perimenopausal symptoms (i.e., menopausal symptom burden) with risk of dementia as determined by both cognitive and behavioural changes. Studies have shown that select perimenopausal symptoms may be related to AD-related neuropathological changes in the brain. For instance, perimenopausal females who experienced night sweats (i.e., a vasomotor symptom) had greater brain amyloid beta pathology than perimenopausal females who did not^61^. While such findings suggest that perimenopausal symptoms may be related to risk of dementia, given that participants in our study reported an average of 3.7 perimenopausal symptoms, it is important to recognize that measuring only one symptom does not tell the whole story; symptomatic perimenopausal females may experience more than one symptom at a time. The experience of multiple menopausal symptoms may estimate dementia risk in a way not accounted for by analysis of a single menopausal symptom, as evidenced by the dose effect we found. Further, to identify whether the experience of multiple menopausal symptoms is related to dementia risk, both markers of cognitive change and behavioural change should be used.

While changes in cognitive function may act as one marker of dementia risk, later-life emergent and persistent changes in behaviour consistent with MBI are another marker, which complements cognition^36^. Several studies have linked MBI with cognitive decline and dementia. A longitudinal study of MBI in cognitively normal older persons, using sex-stratified analyses, described MBI effects for cognitive decline in both females and males^62^. In a mixed sex study that explored progression to dementia in a neuropsychiatry clinic sample, participants with MBI had a higher progression-rate to dementia than those with psychiatric disorders. Additionally, in both female and male patients with MCI, those with comorbid MBI had a higher progression rate than those without^63^. These findings have been replicated in a geriatric psychiatry clinic sample^47^ and in observational cohorts of older adults with normal cognition and subjective cognitive decline^37,64,65^, with MCI^37,38^, and in mixed cognitively unimpaired/MCI samples^42,44–46^. These findings support MBI as an effect modifier in the relationship between cognitive function and incident dementia. Our cross-sectional study shows that this may also be the case in a cohort of females, alone. This emerging body of evidence highlights the importance of including assessment of behavioural changes, consistent with MBI, in conjunction with the well-established practice of determining cognitive status in dementia risk assessment^66^. Despite the importance of the link, sparse research has explored the relationship between menopause and MBI and none exploring dose effects, i.e., number of perimenopausal symptoms and later-life MBI symptoms (let alone cognitive symptoms). Our findings of a dose effect encourage further investigations of this link.

Why some perimenopausal females and not others may experience symptoms has not been elucidated; however, recent evidence suggests that differences in the experience of menopause, such as in menopausal symptoms, may be linked to physiological stressors^10–13^. Estradiol is known to lead to synapse formation, neurite outgrowth, and neurogenesis, all of which decline in neurodegenerative diseases and dementia. However, in menopause, estradiol naturally declines^6,10^. Thus, the experience of menopausal symptoms may act as an indicator of how well females tolerate estradiol changes^16,67–70^. Indeed, in a 10-year longitudinal study of Vitamin D exposure and incident dementia, the effect of Vitamin D for lower hazard of incident dementia was greater in females, thought to be secondary to the role of estrogen in activating Vitamin D^67^. This evidence suggests two possible mechanisms for how menopause may be related to risk for AD. The first mechanism may be the direct effect of estrogens on neurons^17,68^ with greater decline in estrogens increasing risk for cognitive decline and dementia. The second mechanism may be indirect and dependent on how body systems other than the brain in individual females respond to estrogen decline, modifying risk of developing AD-related neuropathological changes in the brain. Although further exploration is required, it is likely that both mechanisms contribute to the association between estradiol loss, perimenopausal symptoms, and AD.

HT are amongst the most effective treatments for alleviating perimenopausal symptoms by mitigating menopause-related estradiol decline^8,69^. Thus, they are essential to understanding physiological responses to menopause. Given that estrogens have been shown to mitigate against AD cognitive and brain changes ^16,68,70–73^, some research speculates they may help to reduce to the risk of dementia^8,16,32,74^. Despite this hypothesis, evidence remains conflicted as studies continue to report mitigating^8,32,33^, neutral^34,35^, and harmful^35,75,76^ associations with dementia risk for females. Additionally, factors such as the age at start of therapy, duration, genetic interactions, and type of HT likely complicate the association, making the relationship highly complex and variable person to person^8,16,33,71,74^.

In our cognitive analysis, we found neither direct nor interaction effects of HT for cognitive function in females independent of menopause type or age at menopause. These findings contradict studies that support the role of HT to ameliorate cognitive decline in females with perimenopausal symptoms ^8,32,33^. This may be due to the highly variable response of individual females to estradiol loss and replacement and the direction of effect in our study does suggest potential enhancement of cognitive performance. Indeed, our behavioural analysis revealed a main effect for HT and behaviour without an interaction effect, suggesting potential links between HT taken during perimenopause and subsequent behavioural risk for dementia in our cohort. Although research remains sparse, other studies show that the use of HT has been implicated in the improvement of behavioural symptoms during perimenopause, including emotional symptoms^75,77–80^. One study found that compared to females who experienced no symptoms of menopause, perimenopausal females who used HT had improved depression severity, but only in those who also experienced hot flashes suggesting that the beneficial effects were due to treatment of the hot flashes^77^. Altogether, these findings suggest that HT may mitigate MBI symptoms differently from cognitive symptoms.

There are several limitations to this study. First, the cross-sectional design of the study limits conclusions about causality. To determine temporal relationships, longitudinal data are required, ideally with biomarker data (i.e., blood estradiol, phosphorylated-tau, and amyloid beta levels) to further explore potential mechanisms. Second, menopausal symptom burden was operationalized objectively as a symptom count, (i.e., number of menopausal symptoms) rather than severity, which might be more subjective. Nonetheless, recent evidence suggests that menopausal symptom severity is associated with poorer cognitive performance^24^. Additional research is required to incorporate severity into symptom count, to explore these relationships further. Third, sample size limitations precluded analyses of differences between types and formulations of HT and progestogens. A growing number of studies suggest that 17β-estradiol HT may exert the greatest of neuroprotective effects on cognitive function and risk for AD as compared to other HT estrogens^81,82^. As we did not find a significant association of HT with cognitive function, it may be that combining different HT estrogens into one group skewed the results. As CAN-PROTECT data accumulate, we may be able to address this limitation in the future. Nonetheless, there are also multiple strengths to this study. The experience of menopause and its association with dementia is highly complex and likely influenced by several variables such as type of menopause, age of onset, and use of HT and progestogens^8,16,33,71,74^. Our study included each of these variables to represent more accurately the experience of menopause. Finally, contributing to the novelty of our study, we included behaviour, operationalized as the validated construct of MBI to assess risk. Together, changes in cognitive function and behaviour represent two important markers for risk of dementia. Therefore, broadening our understanding of how the experience of menopause may be associated with both cognition and behaviour could have clinical applications for risk for incident dementia.

## Conclusions

In conclusion, this study has demonstrated that recalled greater menopausal symptom burden is associated with poorer current cognitive function and greater current MBI symptom burden in mid-to-late life. Use of HT control may help to mitigate MBI symptoms more than cognitive symptoms. These findings suggest menopausal symptom burden may predict susceptibility to dementia, as evidenced by greater cognitive and behavioural factors in those with greater menopausal symptom burden.

## Data Availability

Data cannot be shared publicly as CAN-PROTECT does not yet have a data sharing policy that has been approved by Ethics.

## Acknowledgements

All authors contributed to the study design, data acquisition, data analysis and interpretation, and manuscript revisions. All authors gave approval for the final submission.

## Notes

### Competing Interest Statement

The authors have declared that no competing interests exist.

### Funding Statement

Yes

### Author Declarations

University of Calgary Conjoint Health Research Ethics Board

